# Protocol and Statistical Analysis Plan for the Hyperangulated versus Standard Geometry Laryngoscope Blade (ANGLE) Trial

**DOI:** 10.64898/2026.11.11.26367862

**Authors:** Stephanie C. DeMasi, Matthew W. Semler, Aaron E. Robinson, Heath D. White, Kevin G. Buell, Carolynn Lyle, Michael Gottlieb, LuAnn H. Barnett, Nida Qadir, Amy L. Kloosterboer, Daniel Resnick-Ault, Sheetal Gandotra, Logan L. Beach, Derek J. Vonderhaar, Taya Haas, Matthew J. Barraza, Alexandra June Gordon, Kevin R. Landefeld, Kevin W. Gibbs, Calvin A. Brown, R. Gentry Wilkerson, Stephen C. Lim, Akram Khan, Amelia L. Muhs, Xian Qiao, J. Taylor Herbert, James E. Walker, Jason C. Brainard, David J. Douin, Derek W. Russell, Shekhar A. Ghamande, Karen E. Jackson, Thomas Alcorn, Shannon M. Alwood, Jason Kirincich, Andrew P. Bartuska, Emily K. Bowery, Nicholas Sachs, Micah R. Whitson, Alex P. Brueder, Amy L. Martin, Jonathan C. Henricks, Pranjal B. Gupta, Oscar A. Estrada Paz, Jessica A. Palakshappa, Michael S. Char, Kami M. Hu, David A. Rayburn, Aaron Trimble, Yasin A. Khan, Vijay Krishnamoorthy, Nicholas Raush, Franklin L. Wright, Samuel D. Gilliland, Lauren Daley, Adit A. Ginde, Ariel A. Lewis, Todd W. Rice, Wesley H. Self, Brant Imhoff, Li Wang, Matthew E. Prekker, Brian E. Driver, Jonathan D. Casey, Stacy A. Trent, ANGLE investigators, Pragmatic Critical Care Research Group

**Affiliations:** Department of Emergency Medicine, Vanderbilt Health, Nashville, Tennessee, USA; Vanderbilt Institute for Clinical and Translational Research, Vanderbilt Health, Nashville, Tennessee, USA; Department of Medicine, Division of Allergy, Pulmonary, and Critical Care Medicine, Vanderbilt Health, Nashville, Tennessee, USA; Department of Emergency Medicine, Hennepin County Medical Center, Minneapolis, Minnesota, USA; Department of Medicine, Division of Pulmonary and Critical Care Medicine, Baylor Scott & White Medical Center – Temple, Temple, Texas, USA; Department of Internal Medicine, Division of Pulmonary, Critical Care and Sleep Medicine, Rush University Medical Center, Chicago, Illinois, USA; Department of Emergency Medicine, Denver Health Medical Center, Denver, Colorado, USA; Department of Emergency Medicine, Rush University Medical Center, Chicago, Illinois, USA; Department of Emergency Medicine, Baton Rouge Branch Campus, Louisiana State University Health Sciences Center – New Orleans, New Orleans, Louisiana, USA; Our Lady of the Lake Regional Medical Center, Baton Rouge, Louisiana, USA; Department of Medicine, Division of Pulmonary, Critical Care, and Sleep Medicine, David Geffen School of Medicine at UCLA, Los Angeles, California, USA; Department of Anesthesiology, Perioperative and Pain Medicine, Stanford University School of Medicine, Stanford, California, USA; Department of Emergency Medicine, University of Colorado Anschutz School of Medicine, Aurora, Colorado, USA; Department of Medicine, Division of Pulmonary, Critical Care and Sleep Medicine, University of Maryland School of Medicine, Baltimore, Maryland, USA; Department of Emergency Medicine, University of Alabama at Birmingham Heersink School of Medicine, Birmingham, Alabama, USA; Department of Pulmonary and Critical Care Medicine, Ochsner Health, New Orleans, Louisiana, USA; Department of Respiratory Therapy, Hennepin County Medical Center, Minneapolis, Minnesota, USA; Department of Internal Medicine, Division of Pulmonary, Critical Care, and Sleep Medicine, The Ohio State University, Columbus, Ohio, USA; Department of Emergency Medicine, Division of Emergency Critical Care, Stanford University School of Medicine, Stanford, California, USA; Department of Emergency Medicine, David Geffen School of Medicine at UCLA, Los Angeles, California, USA; Department of Internal Medicine, Section of Pulmonary, Critical Care, Allergy and Immunologic Diseases, Wake Forest University School of Medicine, Winston-Salem, North Carolina, USA; Department of Emergency Medicine, Lahey Clinic and UMass Chan-Lahey School of Medicine, Burlington, Massachusetts, USA; Department of Emergency Medicine, University of Maryland School of Medicine, Baltimore, Maryland, USA; Department of Medicine, Section of Emergency Medicine, Louisiana State University Health Sciences Center – New Orleans, New Orleans, Louisiana, USA; Department of Medicine, Division of Pulmonary, Allergy and Critical Care Medicine, Oregon Health & Science University, Portland, Oregon, USA; Department of Medicine, Section of Pulmonary/Critical Care and Allergy/Immunology, Louisiana State University Health Sciences Center – New Orleans, New Orleans, Louisiana, USA; Sentara Pulmonary, Critical Care & Sleep Specialists, Norfolk, Virginia, USA; Department of Medicine, Section of Pulmonary, Critical Care, & Sleep Medicine, Eastern Virginia Medical School, Macon & Joan Brock Virginia Health Sciences at Old Dominion University, Norfolk, Virginia, USA; Department of Anesthesiology, Division of Critical Care Medicine, Duke University School of Medicine, Durham, North Carolina, USA; Department of Medicine, Baton Rouge Branch Campus, Louisiana State University Health Sciences Center – New Orleans, New Orleans, Louisiana, USA; Department of Anesthesiology, University of Colorado Anschutz School of Medicine, Aurora, Colorado, USA; Department of Medicine, Division of Pulmonary, Allergy, and Critical Care Medicine, University of Alabama at Birmingham Heersink School of Medicine, Birmingham, Alabama, USA; Pulmonary Section, Birmingham VA Medical Center, Birmingham, Alabama, USA; Departments of Emergency Medicine and Medicine, University of Maryland School of Medicine, Baltimore, Maryland, USA; Deming Department of Medicine, Section of Pulmonary Diseases, Critical Care, and Environmental Medicine, Tulane University School of Medicine, New Orleans, Louisiana, USA; Department of Surgery, University of Colorado Anschutz School of Medicine, Aurora, Colorado, USA; Department of Biostatistics, Vanderbilt Health, Nashville, Tennessee, USA; Department of Medicine, Division of Pulmonary, Allergy, and Critical Care Medicine, Hennepin County Medical Center, Minneapolis, Minnesota, USA

## Abstract

**Background:** Emergency tracheal intubation is common and high risk. Although guidelines recommend using a video laryngoscope to intubate critically ill adults, whether a hyperangulated or standard-geometry blade is more likely to achieve successful intubation on the first attempt remains unknown.

**Research Question:** Does the use of a video laryngoscope with a hyperangulated blade increase the incidence of successful intubation on the first attempt among adults undergoing emergency tracheal intubation, compared to use of a video laryngoscope with a standard-geometry blade?

**Study Design and Methods:** The Hyper<u>AN</u>gulated versus Standard <u>G</u>eometry <u>L</u>aryngoscope Blad<u>E</u> (ANGLE) trial is a pragmatic, multicenter, unblinded, parallel-group, randomized trial being conducted in 28 sites (10 emergency departments and 18 intensive care units) in the United States. The trial compares use of a video laryngoscope with a hyperangulated blade versus a standard-geometry blade among 2,500 critically ill adults undergoing emergency tracheal intubation. The primary outcome is successful intubation on the first attempt. The secondary outcome is the incidence of hypoxemia during intubation. Enrollment began on December 30, 2025, and is expected to conclude in 2027.

**Interpretation:** The ANGLE trial will provide important data on the effects of the use of a video laryngoscope with a hyperangulated blade versus standard-geometry blade on successful intubation on the first attempt and other outcomes of emergency tracheal intubation. Specifying the protocol and statistical analysis plan before the conclusion of enrollment increases the rigor, reproducibility, and transparency of the trial.

**Trial Registry:** ClinicalTrials.gov; No.: NCT07223762; URL: www.clinicaltrials.gov

## INTRODUCTION

More than 13 million critically ill adults undergo tracheal intubation annually worldwide.^1,2^ In approximately 10-20% of tracheal intubations in the emergency department (ED) or intensive care unit (ICU), clinicians are unable to intubate the trachea on the first attempt.^3–6^ Failure to intubate on the first attempt during emergency tracheal intubation is associated with an increased risk of hypoxemia, hypotension, and cardiac arrest.^5–8^

The likelihood of successful intubation on the first attempt during emergency tracheal intubation may be affected by the design of the laryngoscope used. Previous data demonstrate that use of a video laryngoscope (a laryngoscope with a camera that displays images of the airway on a screen), rather than a direct laryngoscope (a laryngoscope without a camera or screen), increases the incidence of successful intubation on the first attempt.^3,9^ When using a video laryngoscope, the operator must select a blade—the component inserted into the mouth to displace the tongue and expose the vocal cords. Only two shapes of video laryngoscope blade are routinely used in care of adult patients: hyperangulated and standard-geometry.^3,4,6,10,11^ All major video laryngoscope platforms—including GlideScope, C-MAC, and McGRATH—offer both hyperangulated and standard-geometry blade options.^12–14^

A hyperangulated blade has a pronounced curve designed to follow the tongue and upper airway, creating an angle of approximately 80° between its proximal and distal segments.^15,16^ This curvature is intended to allow visualization of the vocal cords without substantial lifting force or manipulation of the head and neck.^17–21^ A rigid stylet inserted into the endotracheal tube is often used to shape the tube to follow the curvature of the blade. Advancement of the endotracheal tube past the vocal cords and into the trachea is visualized on a video screen because the pronounced curvature generally precludes a direct line of sight from the mouth to the vocal cords.^17^

A standard-geometry blade retains the shape of the traditional Macintosh blade designed for direct laryngoscopy, with an angle of approximately 50° between the proximal and distal segments of the blade.^15,16^ During laryngoscopy, the operator often seats the tip of the blade in the midline vallecula and lifts the handle upward and out to displace the tongue and elevate the epiglottis, allowing visualization of the vocal cords either on the video screen or directly through the mouth.^22^ Compared with a hyperangulated blade, a standard-geometry blade may provide a less complete view of the vocal cords but may facilitate endotracheal tube advancement by creating a more direct path through the oropharynx.^22^ It also permits direct laryngoscopy if the camera view is obscured by blood or emesis.

Whether the shape of the blade affects the incidence of successful intubation on the first attempt or the risk of complications during emergency tracheal intubation remains uncertain.^3,23,24^ To our knowledge, no previous randomized trial has compared use of a hyperangulated blade versus a standard-geometry blade during emergency tracheal intubation of patients in an ED or ICU. Multiple previous trials have compared use of a hyperangulated blade versus a standard-geometry blade among adults undergoing tracheal intubation with a video laryngoscope in the operating room prior to elective surgery, with inconsistent results.^21,25–32^ Most were single-center studies involving small, selected populations, specific brands of laryngoscope, or simulated difficult-airway conditions. The largest previous trial randomized approximately 1,600 patients undergoing intubation before elective surgery to use of a hyperangulated blade or standard-geometry blade; the incidence of successful intubation on the first attempt appeared to be higher in the hyperangulated blade group (87.6%) than in the standard-geometry blade group (82.9%).^25^ However, other studies have suggested that improved glottic visualization with a hyperangulated blade may not translate into higher incidence of successful intubation on the first attempt, with some reporting lower success rates or longer intubation times.^26,32,33^ These findings emphasize that hyperangulated blades may improve glottic visualization and standard-geometry blades may facilitate tube passage, but no randomized trial has yet determined which approach improves overall intubation success for the many critically ill adults who undergo emergency tracheal intubation in an ED or ICU each year.

To address this uncertainty, we designed the Hyper<u>AN</u>gulated versus Standard <u>G</u>eometry <u>L</u>aryngoscope Blad<u>E</u> (ANGLE) trial to compare hyperangulated versus standard-geometry blades among 2,500 adults undergoing emergency tracheal intubation. We hypothesize that use of a hyperangulated blade will increase the incidence of successful intubation on the first attempt compared with use of a standard-geometry blade.

## STUDY DESIGN AND METHODS

This article was written in accordance with Standard Protocol Items: Recommendations for Interventional Trials (SPIRIT) guidelines (Table 1 and Section 2 of e-Appendix 1).^34^

**Table 1:**
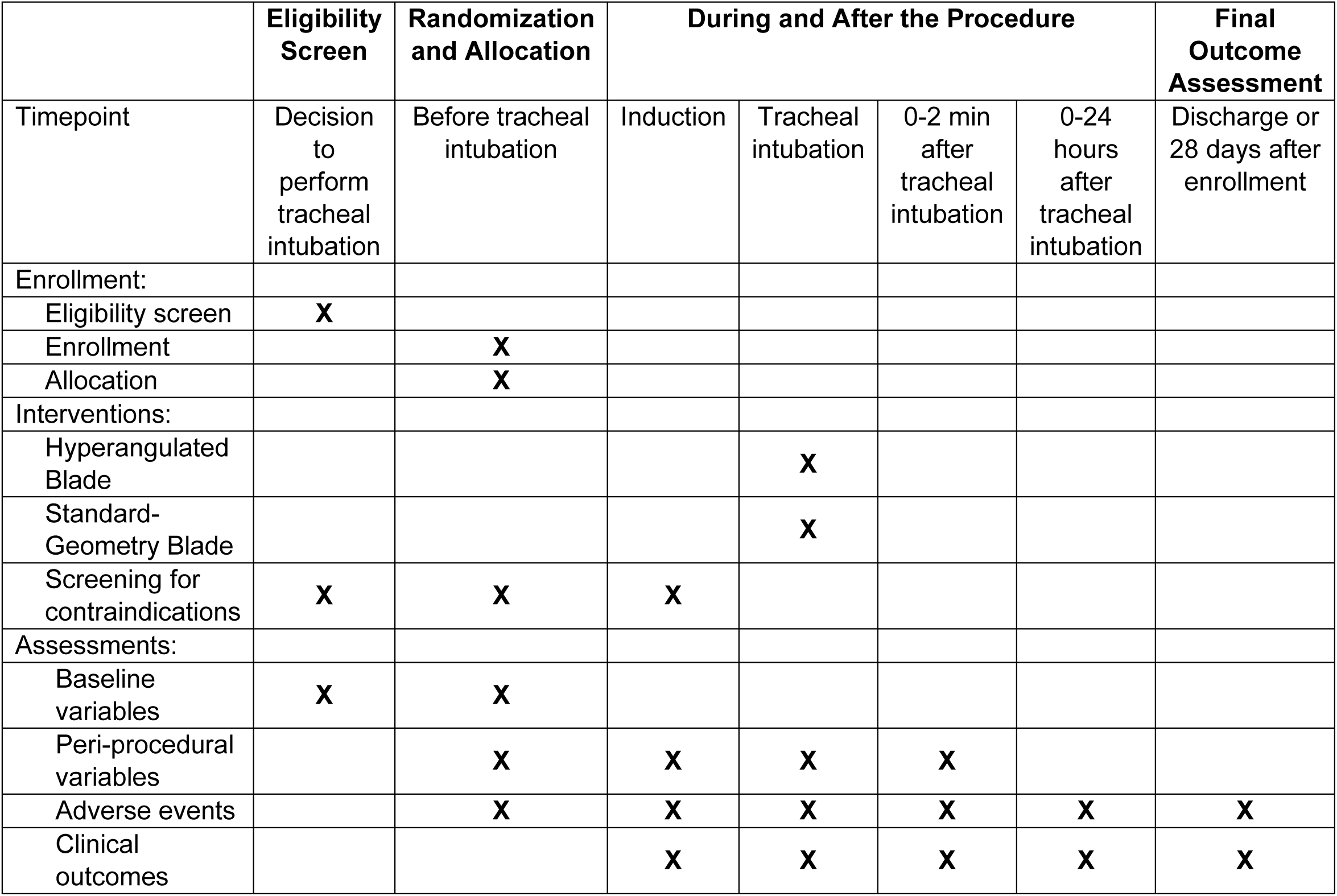
Schedule of Enrollment, Interventions, and Assessments in the ANGLE trial. Table entries without data mean that data elements are not collected at that time.

### Study Design

The ANGLE trial is a pragmatic, multicenter, unblinded, parallel-group, randomized superiority trial comparing the use of a video laryngoscope with a hyperangulated blade versus a standard-geometry blade in 2,500 critically ill adults undergoing emergency tracheal intubation in 28 sites (10 EDs and 18 ICUs) in the United States. The primary outcome is successful intubation on the first attempt. The secondary outcome is the incidence of hypoxemia during intubation. The trial is being conducted by the Pragmatic Critical Care Research Group (www.pragmaticcriticalcare.org). The trial was registered prior to initiation of enrollment (ClinicalTrials.gov identifier: NCT07223762). Information on ethics and regulatory approval and the involvement of patients and the public in trial design and conduct are presented in Sections 3-4 of e-Appendix 1.

### Study Population

Patients undergoing tracheal intubation with a video laryngoscope in a participating ED or ICU are potentially eligible. The trial excludes patients who are known to be pregnant or prisoners. Complete lists of inclusion and exclusion criteria are provided in Table 2.

**Table 2:** Inclusion and Exclusion Criteria.

| **Inclusion Criteria** |
|---|
| Patient is located in a participating unit |
| Planned procedure is orotracheal intubation using a laryngoscope |
| Planned operator is a clinician expected to routinely perform tracheal intubation in the participating unit |
| **Exclusion Criteria** |
| Patient is known to be less than 18 years old |
| Patient is known to be pregnant |
| Patient is known to be a prisoner |
| Immediate need for tracheal intubation precludes safe performance of study procedures |
| Operator has determined that use of a direct laryngoscope is required for the optimal care of the patient |
| Operator has determined that use of a hyperangulated video laryngoscope blade or a standard geometry video laryngoscope blade is required or contraindicated for the optimal care of the patient either because of factors related to the patient (e.g., patient's upper airway anatomy requires a specific blade type) or the operator (e.g., treating clinicians lack sufficient comfort with one of the blade types). |

### Randomization and Treatment Allocation

Patients are randomized in a 1:1 ratio to the use of a hyperangulated blade or a standard-geometry blade in permuted blocks of variable size, stratified by trial site. Trial group assignments are generated using a computerized randomization sequence, placed in sequentially numbered opaque envelopes, and distributed to enrolling sites (Sections 5-6 of e-Appendix 1). Before opening the envelope, the clinician performing the tracheal intubation procedure (referred to as the “operator”) determines that the patient meets eligibility criteria. Trial group assignment remains concealed to patients, trial personnel, and clinicians until after the decision has been made to enroll the patient and the envelope is opened. Patients are enrolled and randomized once the operator opens the trial envelope to reveal trial group assignment. After randomization, patients, clinicians, and trial personnel are not blinded to trial group assignment.

### Study Interventions

#### Hyperangulated Blade Group

For patients assigned to the hyperangulated blade group, clinicians are instructed to use a video laryngoscope with a hyperangulated blade on the first laryngoscopy attempt. The trial considers a blade to be hyperangulated if the blade is designated as hyperangulated by its manufacturer. The trial protocol does not specify the size, manufacturer, brand, or model of hyperangulated video laryngoscope. The models of hyperangulated blades used at the sites participating in the trial are listed in Section 7 in e-Appendix 1 and have an angle between the proximal and distal segments of the blade that ranges from 62.3 to 95.6 (Figure 1).

**Figure 1.**
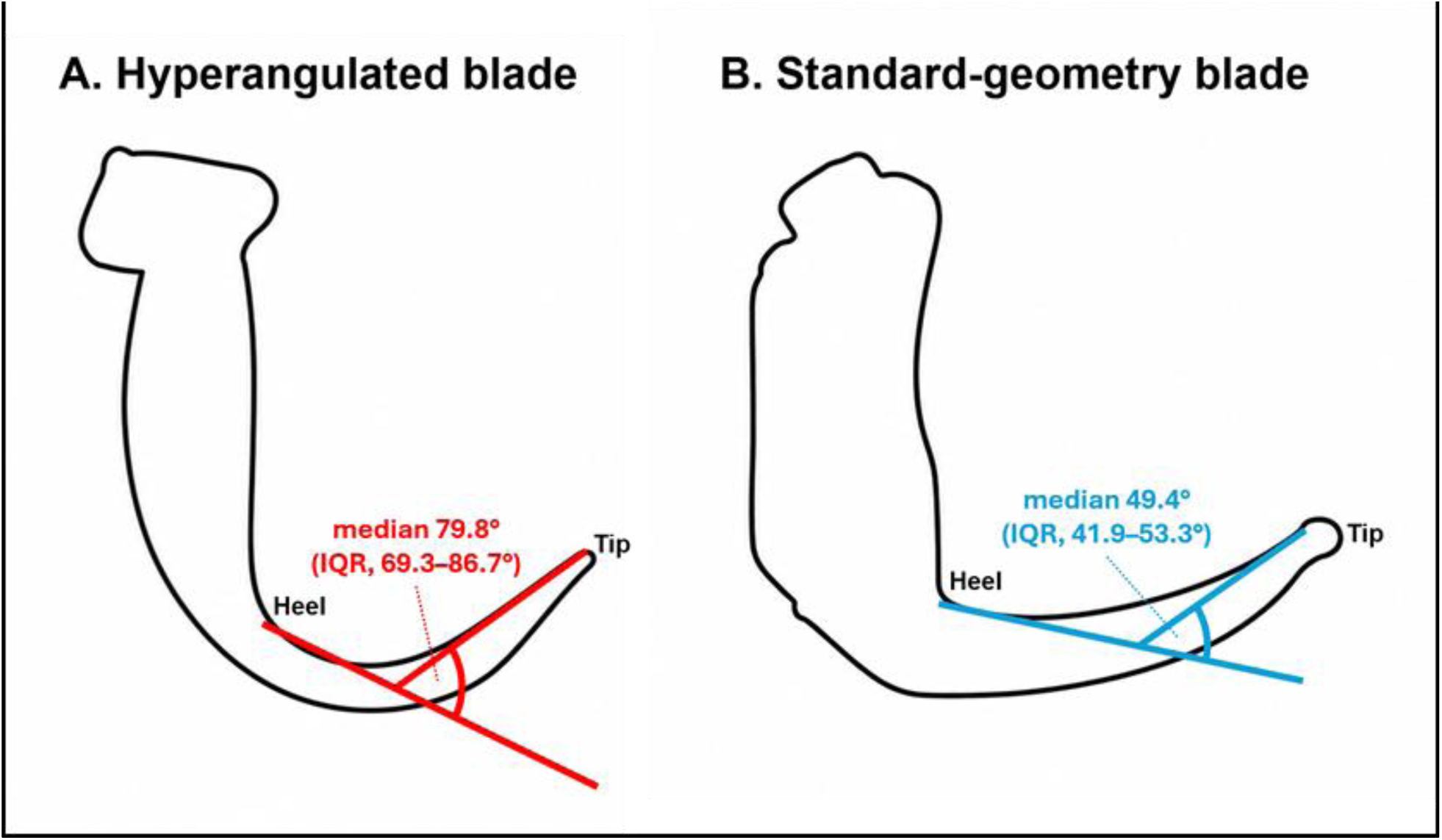
Comparison of Hyperangulated and Standard-Geometry Blades. This figure displays representative outlines of two video laryngoscopes with a hyperangulated blade (A) and a standard-geometry blade (B). The colored lines indicate the directions of the proximal and distal 20-mm segments of the blade’s concave, tongue-facing edge. The arcs mark the angle between these directions for each blade type. This measurement, termed the “heel-to-tip deflection angle,” quantifies intrinsic blade angulation independently of handle orientation. The heel is defined by the intersection of a line along the front of the handle with the blade; the tip is the junction of the distal blade and rounded tip.^16^ Hyperangulated blades commonly available in clinical care have a median angle between the proximal and distal blade segments of 79.8° (interquartile range, 69.3–86.7°), compared with 49.4° (interquartile range, 41.9–53.3°) for standard-geometry blades. The more pronounced curve of a hyperangulated blade is intended to approximate the curvature of the tongue and upper airway and allow visualization of the vocal cords without substantial lifting force or manipulation of the head and neck. In addition to a greater angle between the proximal and distal blade segments, hyperangulated blades have a more deeply bowed shape, smaller average and minimum radii of curvature, and a region of greatest curvature that tends to lie closer to the blade tip than in standard-geometry blades. By comparison, a standard-geometry blade is designed to retain the shape of a traditional Macintosh direct laryngoscope and can be used by the operator to create a direct, line-of-sight path to the vocal cords. The outlines in this example depict a GlideScope Spectrum Hyperangle S3 (A) and a C-MAC Mac 3 (B) and are adapted from Driver BE, Trent SA, DeMasi SC, Robinson AE, Prekker ME. Differentiating standard geometry and hyperangulated video laryngoscope blades by handle-blade angle, blade curvature, and camera field of view. medRxiv [Preprint]. 2026. doi:10.64898/2026.10.22.26366035.

#### Standard-Geometry Blade Group

For patients assigned to the standard-geometry blade group, clinicians are instructed to use a video laryngoscope with a standard-geometry blade on the first laryngoscopy attempt. The trial considers a blade to be standard-geometry if the blade is designated as standard-geometry (or Macintosh) by its manufacturer. The trial protocol does not specify the size, manufacturer, brand, or model of standard-geometry video laryngoscope. The models of standard-geometry blades used at the sites participating in the trial are listed in Section 7 in e-Appendix 1 and have an angle between the proximal and distal segments of the blade that ranges from 40.4 to 56.1 (Figure 1).

### Co-Interventions

The ANGLE trial determines only the type of blade (hyperangulated vs standard-geometry) used on the first laryngoscopy attempt. If the first attempt is unsuccessful, the operator may use any method of intubation on subsequent attempts, including the use of a standard-geometry blade in the hyperangulated group or a hyperangulated blade in the standard-geometry group. If determined to be required to ensure optimal care of the patient, treating clinicians may use any device at any time (including the non-assigned blade type on the first laryngoscopy attempt), regardless of trial group assignment. The type of laryngoscope blade used during the initial and final attempts will be collected and reported.

The trial protocol encourages operators to complete the intubation with use of the video screen (indirect laryngoscopy). The choice between use of a rigid stylet, a malleable stylet, and a bougie to facilitate advancement of the endotracheal tube is at the discretion of treating clinicians. All other aspects of the intubation procedure are at clinicians’ discretion, including selection of sedative and neuromuscular blocking medications, patient positioning, approach to preoxygenation, and size of endotracheal tube. For all patients in the trial, best practices in tracheal intubation are encouraged according to local clinical protocols.

### Data Collection

An observer not directly involved with the intubation procedure collects data in real time for key procedural outcomes, including the number of times a laryngoscope blade, a bougie, and an endotracheal tube enter the patient’s mouth and the lowest oxygen saturation, lowest systolic blood pressure, and administration of vasopressors between induction and two minutes after intubation. Observers are either research personnel or trained clinical personnel (e.g., physicians or nurses). The accuracy of this method of data collection has been validated previously^35^ and used in numerous previous trials of emergency tracheal intubation.^3,11,36^

Immediately after the intubation procedure, the operator completes a paper data collection form to record characteristics of the procedure (e.g., approach to preoxygenation, oxygenation and ventilation between induction and laryngoscopy, laryngoscope and blade used on the first attempt, Cormack-Lehane grade of glottic view,^37^ use of a bougie or a stylet, and equipment used on the final attempt), complications during intubation (e.g., witnessed pulmonary aspiration, cardiac arrest), and the operator’s prior intubating experience, defined as the total number of previous intubations with a direct laryngoscope, total number of previous intubations with a video laryngoscope with a hyperangulated blade, and total number of previous intubations with a video laryngoscope with a standard-geometry blade). Trial personnel at each site review the electronic health record to collect data on baseline characteristics, management before and after laryngoscopy, and in-hospital clinical outcomes.

### Primary Outcome

The primary outcome is successful intubation on the first attempt. Successful intubation on the first attempt is defined as placement of an endotracheal tube in the trachea with a single insertion of a laryngoscope blade into the mouth and *either* a single insertion of an endotracheal tube into the mouth *or* a single insertion of a bougie into the mouth followed by a single insertion of an endotracheal tube over the bougie into the mouth.

Data for the assessment of the primary outcome are collected independently (i) by an observer trained in recording the number of insertions of the laryngoscope, bougie, and endotracheal tube into the mouth and (ii) by the operator’s self-report following the procedure. Cases with disagreement between the observer and operator are classified as not representing successful intubation on the first attempt. In the event that data from the independent observer are missing, data from the operator’s self-report of successful intubation on the first attempt will be used.

### Secondary Outcome

The sole secondary outcome is hypoxemia, defined as a peripheral oxygen saturation of < 85% during the interval between induction and 2 minutes after tracheal intubation.

### Procedural, Clinical, and Safety Outcomes

Table 3 reports the exploratory procedural, safety, and clinical outcomes. Additional information on outcome definitions is in Section 8 of e-Appendix 1.^3,11^ Information on collection, assessment, and reporting of Adverse Events is in Section 9 in e-Appendix 1.

**Table 3:** Trial outcomes.

|  |  |
|---|---|
| **Primary Outcome** | Successful intubation on the first attempt, defined as placement of an endotracheal tube in the trachea with a single insertion of a laryngoscope blade into the mouth and EITHER a single insertion of an endotracheal tube into the mouth OR a single insertion of a bougie into the mouth followed by a single insertion of an endotracheal tube over the bougie into the mouth. |
| **Secondary Outcome** | Hypoxemia, defined as a peripheral oxygen saturation < 85% during the interval between induction and 2 minutes after tracheal intubation. |
| **Exploratory Procedural Outcomes** | Cormack-Lehane grade of glottic view on the first laryngoscopy attempt |
|  | Number of laryngoscopy attempts |
|  | Number of bougie attempts |
|  | Number of endotracheal tube attempts |
|  | Duration of intubation, defined as the time (in seconds) between the initial insertion of the laryngoscope blade into the mouth and the final placement of an endotracheal tube or tracheostomy tube in the trachea |
|  | Severe complications during intubation, defined as the development of one or more of the following between induction and 2 minutes after intubation: (1) severe hypoxemia (SpO2 <80%), (2) severe hypotension (systolic blood pressure <80 mm Hg or new or increased vasopressor administration), and (3) cardiac arrest |
| **Exploratory Safety Outcomes** | Esophageal intubation |
|  | Injury to the teeth |
|  | Operator-reported aspiration |
| **Exploratory Clinical Outcomes** | In-hospital death by 1 hour |
|  | In-hospital death by 28 days |
|  | Ventilator-free days through day 28 |
|  | ICU-free days through day 28 |

### Sample Size Estimation

The ANGLE trial was designed to detect a 5-percentage-point absolute difference between groups in the incidence of successful intubation on the first attempt. A difference of this magnitude is similar to or smaller (more conservative) than that considered clinically meaningful in prior airway management trials.^3,10,38^ Because clinicians choose between these blade geometries for millions of emergency tracheal intubations each year, a high degree of certainty regarding their comparative effectiveness was considered essential; therefore, the trial was designed to provide 95% statistical power. Based on a recent trial conducted in the same ED and ICU settings, we assumed that successful intubation on the first attempt would occur in 85% of patients in the standard-geometry blade group.^3^ We calculated that achieving 95% statistical power at a 2-sided α of 0.05 to detect a 5-percentage-point absolute increase—from 85% in the standard-geometry blade group to 90% in the hyperangulated blade group—would require enrollment of 2,350 patients (1,175 per group). To allow for missing primary-outcome data in up to 5% of patients, planned enrollment was increased to 2,500 patients (1,250 per group).

### Data and Safety Monitoring Board

The composition and responsibilities of the independent Data and Safety Monitoring Board (DSMB) are described in Section 10 of e-Appendix 1. The DSMB developed a charter, approved the trial protocol, and approved the trial monitoring plan prior to initiation of enrollment. The DSMB has the authority to recommend that the trial stop at any point, request additional data, request additional interim analyses, or request modifications to the trial protocol.

### Interim Analysis

On October 23, 2026, the DSMB reviewed a single interim analysis, prepared by the unblinded trial biostatistician at the anticipated halfway point of the trial after enrollment of 1,250 patients. The prespecified stopping boundary for efficacy was a P-value ≤ 0.001 for the difference between trial groups in the primary outcome (successful intubation on the first attempt) using a Chi-square test. This conservative Haybittle–Peto boundary will allow the final analysis to be performed using an unchanged level of significance (two-sided P-value < 0.05). After reviewing the interim analysis, the DSMB recommended continuing the ANGLE trial without modification.

### Statistical Analysis Principles

Analyses will be conducted following reproducible research principles using R software (R Foundation for Statistical Computing, Vienna, Austria). Categorical variables will be presented as number and percentage. Continuous variables will be presented with mean and SD or median and IQR, as appropriate. A two-sided P-value of < 0.05 will define a statistically significant between-group difference in the primary outcome. With a single primary outcome, no adjustment for multiplicity will be made. For analyses of secondary and exploratory outcomes, emphasis will be placed on the magnitude of differences between groups with 95% confidence intervals (which will not be adjusted for multiple comparisons), rather than statistical significance. P-values will not be presented.

### Main Analysis of the Primary Outcome

The primary analysis will be an intention-to-treat comparison of patients randomized to the hyperangulated blade group versus the standard-geometry blade group with regard to the primary outcome of successful intubation on the first attempt. The difference between groups will be examined using an unadjusted Chi-square test without Yates’ continuity correction. The absolute risk difference, associated 95% confidence interval, and P-value for the comparison will be presented. Additional analyses of the primary outcome are described in Section 11 of e-Appendix 1.

### Analyses of heterogeneity of treatment effect

Heterogeneity of treatment effect is nonrandom variation in the magnitude or direction of the effect of a treatment on an outcome across levels of a baseline covariate. Our subgroup analyses will examine whether prespecified baseline variables modify the effect of trial group assignment (hyperangulated blade vs standard-geometry blade) on the primary outcome using a formal test of statistical interaction in a logistic regression model with the primary outcome as the dependent variable and trial group, the proposed effect modifier, and the interaction between trial group and the proposed effect modifier included as independent variables. In accordance with the Instrument for assessing the Credibility of Effect Modification Analyses (ICEMAN) recommendations,^39^ we have prespecified the following limited number of baseline variables as potential effect modifiers and provided details of the hypothesized direction of effect modification for each effect modifier in Section 12 of e-Appendix 1:

1. Body-mass index (kg/m^2^)
2. Patient location (ED / ICU)
3. Trauma (Yes / No)
4. Cervical Spine Immobilization (Yes / No)
5. Operator’s pre-enrollment assessment of the anticipated difficulty of intubation (Easy, Moderate-or-difficult, Not Recorded)
6. Operator’s pre-enrollment assessment of the presence of difficult airway characteristics (Section 13 of e-Appendix 1) (Present, Absent, Not Recorded)
7. Operator experience at the time of enrollment (Section 14 of e-Appendix 1)

a. Total number of operator’s previous intubations
b. Proportion of operator’s previous intubations performed using a hyperangulated blade

### Analysis of the Secondary Outcome

We will perform an unadjusted, intention-to-treat comparison of patients randomized to the hyperangulated blade group versus the standard-geometry blade group with regard to the secondary outcome of hypoxemia. We will report the absolute difference in proportions and the corresponding 95% confidence interval calculated using the Wald method.^40^

### Analyses of Additional Outcomes

We will conduct unadjusted, intention-to-treat analyses comparing patients randomized to the hyperangulated blade group versus the standard-geometry blade group with regard to each prespecified exploratory procedural outcome, exploratory clinical outcome, and safety outcome. Between-group differences and the associated 95% confidence intervals will be generated using the Wald method for categorical outcomes and a Wilcoxon rank-sum test for continuous outcomes.

### Handling of Missing Data

We anticipate that no data on the primary outcome will be missing except for patients who choose to withdraw their data or are withdrawn from the trial after being discovered to have been prisoners at the time of intubation. When data are missing for an outcome, we will perform complete-case analysis, excluding cases where the data for the analyzed outcome are missing.

In adjusted analyses, missing data for covariates will be imputed using multiple imputation. All variables included in prespecified statistical models will also be included in the imputation model. This allows for the relationships held between the variables of interest to be as similar as possible with and without imputation. The method of multiple imputation that will be used in adjusted analyses uses bootstrapping and predictive mean matching. Samples of non-missing data will be bootstrapped, then fit to an additive regression model to predict missing data. Imputation will be performed using the R Statistical software’s aregImpute function from the Hmisc package.

### Trial Status, Protocol Changes, and Data Sharing

Information on the status of the trial, plans for documenting future protocol changes, and privacy, data storage, and data sharing are in Sections 15-17 of e-Appendix 1.

## Discussion

This manuscript reports the rationale, design, and analysis plan for the ANGLE trial, a 2,500-patient randomized trial evaluating the effect of use of a video laryngoscope with a hyperangulated blade versus a standard-geometry blade among adults undergoing emergency tracheal intubation. The trial is designed to estimate the effect of blade geometry across the devices, clinical practices, patients, and operators encountered during emergency tracheal intubation in EDs and ICUs. Several design choices support this objective.

The ANGLE trial allows clinicians to use any available video laryngoscope platform and any model of blade within the assigned geometry class (hyperangulated vs standard-geometry). This design addresses the fundamental question of the effect of blade shape on outcomes, rather than a narrower question about a particular platform or model. All major video laryngoscope platforms, including GlideScope, C-MAC, and McGRATH, now offer both hyperangulated and standard-geometry blades. Across manufacturers and blade models, hyperangulated blades consistently have greater intrinsic blade angulation than standard-geometry blades.^16^ Each hyperangulated blade is more similar in intrinsic blade angulation to hyperangulated blades from different manufacturers than to standard-geometry blades from the same manufacturer.^16^ The ANGLE trial aims to inform clinicians’ choice between hyperangulated and standard-geometry blades regardless of the specific video laryngoscope platform and models of blade available in their practice.

The ANGLE trial defers the choice of which intubation adjunct to use with the assigned video laryngoscope blade—a rigid stylet, malleable stylet, or bougie—to the treating clinician. A prior multicenter trial comparing a bougie with a malleable stylet during emergency tracheal intubation with a standard-geometry blade reported similar rates of successful intubation on the first attempt.^38^ Although some experts recommend that a hyperangulated blade always be paired with a rigid stylet, a trial in the operating room reported an incidence of successful intubation on the first attempt with hyperangulated video laryngoscope blades that was at least as high with a bougie as with a rigid stylet.^41^ Therefore, the choice between adjuncts was left to the discretion of clinicians.

Successful intubation on the first attempt, the primary outcome of the ANGLE trial, is the most commonly used endpoint in trials evaluating the performance of emergency tracheal intubation.^3,10,38^ In observational research, failure to intubate on the first attempt is associated with higher incidences of hypoxemia, hypotension, cricothyroidotomy, and cardiac arrest.^5,6,42^ Two prior randomized trials found that video laryngoscopy increased the incidence of successful intubation on the first attempt compared with direct laryngoscopy without demonstrating fewer complications; however, both trials had limited power to detect differences in complications.^3,43^ Meta-analyses suggest that video laryngoscopy, by increasing the incidence of successful intubation on the first attempt, may decrease the incidence of hypoxemia.^44^

An important consideration for trials evaluating equipment and procedural techniques is the prior experience of the clinician performing the procedure. The effect of blade geometry on outcomes could depend on an operator’s overall number of previous intubations or the proportion of an operator’s previous intubations that were performed with that specific blade geometry. Effect modification by operators’ previous experience and variation in operator populations between studies could help explain the heterogeneity in the results in prior studies. Prior studies generally have been limited to small and homogeneous populations of operators with either limited or extensive prior intubating experience^19,21,29^ or significantly more previous experience with one blade geometry than the other.^26^ To address the role of operators’ previous experience, the ANGLE trial deliberately selected sites that represent the full range of overall experience levels (trainees conducting their first intubation to highly experienced attending physicians and nurse anesthetists) and blade-specific experience levels (operators whose experience is primarily with hyperangulated blades, operators whose experience is primarily with standard-geometry blades, and operators with a similar amount of prior experience using each blade geometry). By including hundreds of operators with a broad range of prior experience, collecting detailed data on operator experience (overall and with each blade geometry), and prespecifying analyses of effect modification by operator experience (overall and with each blade geometry), the ANGLE trial will be able to rigorously quantify the effect of blade geometry overall and examine if the effect is different for specific groups of operators.

## Data Availability

All data produced in the present study may be available upon reasonable request to the authors

## Interpretation

The ANGLE trial will provide important evidence regarding the effect of use of a hyperangulated blade compared with use of a standard-geometry blade on the incidence of successful intubation on the first attempt among critically ill adults undergoing emergency tracheal intubation with a video laryngoscope. To aid in the transparency and interpretation of trial results, this article detailing the protocol and statistical analysis plan for the ANGLE trial was finalized prior to the conclusion of patient enrollment.

## Author Contribution

Study concept and design: S.C.D, M.W.S, M.E.P, B.E.D, J.D.C, S.A.T; Drafting of the manuscript: S.C.D, M.W.S, J.D.C, S.A.T; Critical revision of the manuscript for important intellectual content: all authors.

## Acknowledgments

A full list of authors and collaborators are listed in the supplement.

## Disclaimer

None

## Declaration of generative AI and AI-assisted technologies in the manuscript preparation process

The authors wrote the complete initial draft of the manuscript. During revisions, the authors used ChatGPT (OpenAI, GPT-6 and Copilot) to review the text and references for accuracy and clarity. Revisions were implemented by the authors who take full responsibility for the accuracy, integrity, and final manuscript content.

