## Supplementary material for "Protocol and Statistical Analysis Plan for the Hyperangulated versus Standard Geometry Laryngoscope Blade (ANGLE) Trial": e-Appendix 1


1. List of ANGLE Investigators 2
2. SPIRIT 2013 Checklist 7
3. Ethics and Regulatory Approval 13
4. Patient and Public Involvement 15
5. Group Assignment Sheet (Hyperangulated blade group) 17
6. Group Assignment Sheet (Standard-geometry blade group) 18
7. Angle Between the Proximal and Distal Blade Segments of Hyperangulated vs Standard-Geometry Video Laryngoscope Blades 19
8. Definitions of Free-Day Outcomes 20
9. Safety Monitoring and Adverse Events 21
    9.1 Adverse Event Definitions 21
    9.2 Monitoring for Adverse Events 23
    9.3 Recording and Reporting Adverse Events 24
    9.4 Clinical Outcomes that may be Exempt from Adverse Event Recording and Reporting 25
10. Composition and Responsibilities of the Data and Safety Monitoring Board 27
11. Additional Analyses of the Primary Outcome 28
12. Analyses of heterogeneity of treatment effect 29
13. Rationale for the approach to collecting, reporting, and analyzing difficult airway characteristics and anticipated difficulty of intubation 33
14. Rationale for the approach to collecting, reporting, and analyzing operator experience 36
15. Plan for Communication of Protocol Changes 37
16. Patient Privacy, Data Storage, and Sharing 38
17. Trial Status 39
18. References 40

#### 1. List of ANGLE Investigators

Coordinating Center: *Clinical Coordinating Center (Vanderbilt University Medical Center, Nashville, Tennessee)* – Jonathan D. Casey, MD, MSc* (Co-Director); Matthew W. Semler, MD, MSc* (Co-Director); Ariel A. Lewis, MPH, BSN, RN* (Project Management Lead); Stephanie C. DeMasi, MD, MSc* (Airway Trials Program Director); Todd A. Hendrix, RRT; Bradley D. Lloyd, RRT-ACCS; Karen F. Miller, RN, MPA; Tracy L. Morgan, RRT; Michele Nguyen, MSPH; Kevin P. Seitz, MD, MSc; Todd W. Rice, MD, MSc*; Wesley H. Self, MD, MPH*; Kaitlyn R. Whitaker, MS; LaKeysha C. Wiggins. *Data Coordinating Center (Vanderbilt University Medical Center, Nashville, Tennessee)* – Brant Imhoff, MS*; Matthew S. Shotwell, PhD; Li Wang, MS*.

Baylor Scott and White Medical Center – Temple: Heath D. White, DO, MS*; Shekhar A. Ghamande, MD*; Tayler J Acton, DO; Christian Beltran, MD

Denver Health Medical Center: Stacy A. Trent, MD, MPH*; Carolynn Lyle, PA-C, MPH*; Sophia Rocha Ellerby, MPH; Madison Stiglich, MPH

Duke University School of Medicine: J. Taylor Herbert, MD, PhD*; Vijay Krishnamoorthy, MD, PhD, MPH*; Vincent Ciampi, PA, RRT-ACCS, RCP; Eric Conklin, MD; John Levi, MD; Ibtehaj Naqvi, MD, PhD; Richard T. Young, DO

Hennepin County Medical Center: Brian E. Driver, MD*; Taya Haas, MBA, RRT*; Amy L. Martin, BS, RRT*; Andrew P. Bartuska MD*; Matthew E. Prekker, MD, MPH*; Aaron E. Robinson, MD, MPH*; Salma Awale, BS; Meghana Chimata, BS, BA; Chiara Costamagna Soto; Clara Duray; Laurynn Giles, BS; Salma Hassan, BA; Franciska Hauer, BS, BA;

Audrey Hendrickson, MPH; Kowsar Hurreh, BA; Pang Khang, BS; Brandon Kim, BA; Joanna Kuo, BS; Ruthvika Murikinati, MS; Robert Murphy, BS; Ariam Mussiel, BA; Julianna Prohofsky, BS; Shawna Ratanpal, BS; Simren Samba, BS

Lahey Clinic: Calvin A. Brown III, MD*; Michael S. Char, MD*; Jiwon Chung; Caroline Schissel

Louisiana State University School of Medicine: Yasin A. Khan, MD, PhD*; Stephen C. Lim, MD*; Amelia L. Muhs, MD*; David A. Rayburn, MD*

The Ohio State University: Matthew J. Barraza, MD, MS*; Jonathan C. Henricks, MD*; Christina L. Beran, PharmD; Matthew C. Exline, MD, MPH; Shilpa Sridhar, MD

Oregon Health & Science University: Akram Khan, MBBS*; Aaron Trimble, MD*; Genesis Briceno; Ben Bui; Zoha Imran; Riya Mathew; Jose Pena; Edvinas Pocius; Emily Tribbett

Ochsner Medical Center: Derek J. Vonderhaar, MD*; Alex P. Brueder, MD*; Julie Castex, MSN, APRN, ACNS-BC; Amanda Hayden; Rosemary Liberto, MPH, CCRC; Jainika Savani, MPH

Our Lady of the Lake Medical Center: LuAnn H. Barnett, MD*; Shannon M. Alwood, MD*; James Walker, MD, PhD*; Nicholas Raush, MD*; Connor Bennett, DO; John R. Campbell, MD; Benjamin C. Creel, MD; Greggory R. Davis, PhD; Mary E. Duhon, RN, APNP – CNP; Tina M. Martinez; Melody Salmanian, MD

Rush University Medical Center: Kevin G. Buell, MBBS, MS*; Michael Gottlieb, MD, MBA*; Thomas Alcorn, MD*; Karen E. Jackson, MD*; Amanda Apato, PharmD; Wilfredo Cruz Ayala, MS; Hannah Frank, PharmD; Kristyn Gatling, MA; Stephany Nunez Cruz, PharmD; Michelle Santangelo, MS; Giles Slocum, PharmD; Christopher Gore, MSc; Nirali Patel, MBA; Kyle Damon, BSBA; Ryan Schertz, MBA; Ashley Lachowicz, MSc

Sentara Healthcare: Xian Qiao, MD*; Elizabeth Batchelor, MD; Matthew Bernens, MD; David Gakhokidze, MD; Mahvish Haider, DO; Reece Hoerle, MD; Jessica Kaushal, MBBS; Yuliya Levkiavska, MD; Ginny Marmolejos, MD; Emory Newkirk, MD; Sanjana Ramakrishnan, MD; Azeem Rathore, DO; Rutul Shah, MBBS; Anna Shannon, RN, BSN; Ahmadur Siddiqui, MBBS; Joshua Sill, MD; Zack Soheim, MD; Amanda Cecchini, MD

Stanford University School of Medicine: Alexandra June Gordon, MD*; Amy L. Kloosterboer, MD*; Pranjal B. Gupta, MD*; Andrew P. Bartuska, MD*; Terence Ahern, MD; Leonard Basobas, MS; Samantha Ferguson, BS; Vikram Fielding-Singh, MD, JD; Rana Kabeer, MD, MPH, MBA; Della Lau, BSN, RN; Lilian (Lily) Lau, MHS; Paige Lelati, PA-C; David L. Stahl, MD; Grace Kyin-Ye Tam, BS; Alexis Touros, BS; Melissa A. Vogelsong, MD; Jennifer G. Wilson, MD, MS

University of Alabama at Birmingham Medical Center and Heersink School of Medicine: Logan L. Beach, MD*; Lauren Daley, MD*; Derek W. Russell, MD*; Micah R. Whitson, MD*; Jarad Anderson, MD; Whitney Gulledge, MD; Tyler Power, MD; Callie O'Bryant, MD; Zach Jasper, MD; Miller Cone, MD; Nick Orris, MD; Liz Pittman, MD; Alyson Ridgeway, MD; Richard Preus, MD; Charles Khoury, MD; Jaron Raper, MD; Julie Brown, MD; Jason

Brown, MD; Erin Shufflebarger, MD; Matt Kelly, MD; Todd Peterson, MD; Joel Evans, MD; Kat Griesmer, MD; Luke Burleson, MD

University of California Los Angeles School of Medicine: Kevin R. Landefeld, MD*; Nida Qadir, MD*; Oscar A. Estrada Paz, MD*; Jason Kirincich, MD*; Maria F. Balbuena Bojorquez; Joanne M. Bando, MD; Fardean Khajehmiraki; Sam Limsuwannarot; Elizabeth A. Stone; Julia Vargas; Sherry Xu, RN; Timothy J. Young, MD

University of Colorado School of Medicine: Daniel Resnick-Ault, MD*; David J. Douin, MD, MSc*; Jason C. Brainard, MD*; Emily K. Bowery, BS, BA*; Samuel D. Gilliland, MD*; Adit A. Ginde, MD, MPH*; Franklin L. Wright, MD*; Sarah M. Alber, MD; Tristenne L. Cranford, BS; Yvette R. Evans, MA, BA; Carlee Harris, BSN, RN, CCRN; Emily G. Helmer, BA; Nora M. Houseman, BS; Stephanie G. Martinez, MSc; Catherine D. Mickey, MSN, RN, CCRN-CSC; Jenna S. Naslund, MSN, APRN, ACCNS-AG, CCRN, TCRN; Kyle Pickard, BS; Rana Ramadan, MD; Benjamin K. Scott, MD; Anabelle G. Stevens, BS; Nicholas R. Teman, MD

University of Maryland School of Medicine: Sheetal Gandotra, MD*; R. Gentry Wilkerson, MD*; Kami M. Hu, MD*; Nicholas Sachs, MD, MPH*; Mohammad Abdulelah; Jessica Brenner, MSN, RN; Cihan Caglayan, MD; Angela Calahatian; Caleb Chan, MD, MPH; Emmylou Cui, MS, BSN, RN; Ashli Everstine, MD; Cole Fryman, MD; Eileen Girling, MSN, RN, CCRN; Allison Hanlon, BSN, RN, CCRN; Alena Hershfeld, MS; Ashley Hood, MSN, RN; Daniel Janka, MD; Lucia Joseph, MD; Michael Kenski, BSN, RN, CCRN; Kendall E. Klein, MD; Olga Kolesnik, MD; Allison LaRocco, MD; Soowon Lee; Tanner Long, MD; Martha Lopez Siles, MD; Kristyn McLeod, MD; Heather Murphy, MHA, RN, CCRN; Kalila Murray, BSN, RN, CCRN; Andrew Piner, MD; Lynnee Roane, PhD, MS, RN; Valeriya

Rodionova, MD; Savannah Vogel, MD; Diane Y. Wang, MD; Zachary Wynne, MD; Kevin Yin, BS

Wake Forest Medical Center: Kevin W. Gibbs, MD*; Jessica A. Palakshappa, MD, MS*; Jacqueline M. Cain, BS; Rebecca Edmonds, RN; Paul Finklestein, RN; Copeland Graham, RN; Spencer Harwood, RN; Emery Johnson, RN, MPH; Charlotte Martin, BA; Alicia Money, RN; Benjamin Richards, RN; Matthew Strong, RN; Charles Yarbough, RN; Darija Ward, BS, MBA

*Denotes members of the writing committee who are listed as authors on the manuscript. The remainder of the ANGLE investigators represent collaborators.

STANDARD PROTOCOL ITEMS: RECOMMENDATIONS FOR INTERVENTIONAL TRIALS

#### 2. SPIRIT 2013 Checklist

SPIRIT 2013 Checklist: Recommended items to address in a clinical trial protocol and related documents*

| Section/item | Item No | Description | Addressed on page number |
|---|---|---|---|
| **Administrative information** | | | |
| Title | 1 | Descriptive title identifying the study design, population, interventions, and, if applicable, trial acronym | 1 |
| Trial registration | 2a | Trial identifier and registry name. If not yet registered, name of intended registry | 2 |
| | 2b | All items from the World Health Organization Trial Registration Data Set | 1-14, title page |
| Protocol version | 3 | Date and version identifier | NA |
| Funding | 4 | Sources and types of financial, material, and other support | Title page |
| Roles and responsibilities | 5a | Names, affiliations, and roles of protocol contributors | Title page |
| | 5b | Name and contact information for the trial sponsor | Title page |
| | 5c | Role of study sponsor and funders, if any, in study design; collection, management, analysis, and interpretation of data; writing of the report; and the decision to submit the report for publication, including whether they will have ultimate authority over any of these activities | Title page |

| | | | |
|---|---|---|---|
| | 5d | Composition, roles, and responsibilities of the coordinating centre, steering committee, endpoint adjudication committee, data management team, and other individuals or groups overseeing the trial, if applicable (see Item 21a for data monitoring committee) | Supplement Sections 1, 10 |
| **Introduction** | | | |
| Background and rationale | 6a | Description of research question and justification for undertaking the trial, including summary of relevant studies (published and unpublished) examining benefits and harms for each intervention | 3-5 |
| | 6b | Explanation for choice of comparators | 4-5, 7-8 |
| Objectives | 7 | Specific objectives or hypotheses | 5 |
| Trial design | 8 | Description of trial design including type of trial (eg, parallel group, crossover, factorial, single group), allocation ratio, and framework (eg, superiority, equivalence, noninferiority, exploratory) | 6 |
| **Methods: Participants, interventions, and outcomes** | | | |
| Study setting | 9 | Description of study settings (eg, community clinic, academic hospital) and list of countries where data will be collected. Reference to where list of study sites can be obtained | 6, Supplement Section 1 |
| Eligibility criteria | 10 | Inclusion and exclusion criteria for participants. If applicable, eligibility criteria for study centres and individuals who will perform the interventions (eg, surgeons, psychotherapists) | Table 2 |
| Interventions | 11a | Interventions for each group with sufficient detail to allow replication, including how and when they will be administered | 7-8, Figure 1, Supplement Sections 5-7 |
| | 11b | Criteria for discontinuing or modifying allocated interventions for a given trial participant (eg, drug dose change in response to harms, participant request, or improving/worsening disease) | 8 |
| | 11c | Strategies to improve adherence to intervention protocols, and any procedures for monitoring adherence (eg, drug tablet return, laboratory tests) | Supplement Sections 5, 6 |
| | 11d | Relevant concomitant care and interventions that are permitted or prohibited during the trial | 8 |

| | | | |
|---|---|---|---|
| Outcomes | 12 | Primary, secondary, and other outcomes, including the specific measurement variable (eg, systolic blood pressure), analysis metric (eg, change from baseline, final value, time to event), method of aggregation (eg, median, proportion), and time point for each outcome. Explanation of the clinical relevance of chosen efficacy and harm outcomes is strongly recommended | 10-11, Table 3 |
| Participant timeline | 13 | Time schedule of enrollment, interventions (including any run-ins and washouts), assessments, and visits for participants. A schematic diagram is highly recommended (see Figure) | Table 1 |
| Sample size | 14 | Estimated number of participants needed to achieve study objectives and how it was determined, including clinical and statistical assumptions supporting any sample size calculations | 11 |
| Recruitment | 15 | Strategies for achieving adequate participant enrollment to reach target sample size | NA |
| **Methods: Assignment of interventions (for controlled trials)** | | | |
| Allocation: | | | |
| Sequence generation | 16a | Method of generating the allocation sequence (eg, computer-generated random numbers), and list of any factors for stratification. To reduce predictability of a random sequence, details of any planned restriction (eg, blocking) should be provided in a separate document that is unavailable to those who enroll participants or assign interventions | 6-7 |
| Allocation concealment mechanism | 16b | Mechanism of implementing the allocation sequence (eg, central telephone; sequentially numbered, opaque, sealed envelopes), describing any steps to conceal the sequence until interventions are assigned | 6-7 |
| Implementation | 16c | Who will generate the allocation sequence, who will enroll participants, and who will assign participants to interventions | 6-7 |
| Blinding (masking) | 17a | Who will be blinded after assignment to interventions (eg, trial participants, care providers, outcome assessors, data analysts), and how | 6-7 |
| | 17b | If blinded, circumstances under which unblinding is permissible, and procedure for revealing a participant’s allocated intervention during the trial | NA |
| **Methods: Data collection, management, and analysis** | | | |

| | | | |
|---|---|---|---|
| Data collection methods | 18a | Plans for assessment and collection of outcome, baseline, and other trial data, including any related processes to promote data quality (eg, duplicate measurements, training of assessors) and a description of study instruments (eg, questionnaires, laboratory tests) along with their reliability and validity, if known. Reference to where data collection forms can be found, if not in the protocol | 9 |
| | 18b | Plans to promote participant retention and complete follow-up, including list of any outcome data to be collected for participants who discontinue or deviate from intervention protocols | 9, 15 |
| Data management | 19 | Plans for data entry, coding, security, and storage, including any related processes to promote data quality (eg, double data entry; range checks for data values). Reference to where details of data management procedures can be found, if not in the protocol | Supplement Section 16 |
| Statistical methods | 20a | Statistical methods for analysing primary and secondary outcomes. Reference to where other details of the statistical analysis plan can be found, if not in the protocol | 13-15 |
| | 20b | Methods for any additional analyses (eg, subgroup and adjusted analyses) | Supplement Sections 11, 12 |
| | 20c | Definition of analysis population relating to protocol non-adherence (eg, as randomised analysis), and any statistical methods to handle missing data (eg, multiple imputation) | 13-15 |
| **Methods: Monitoring** | | | |
| Data monitoring | 21a | Composition of data monitoring committee (DMC); summary of its role and reporting structure; statement of whether it is independent from the sponsor and competing interests; and reference to where further details about its charter can be found, if not in the protocol. Alternatively, an explanation of why a DMC is not needed | Supplement Section 10 |
| | 21b | Description of any interim analyses and stopping guidelines, including who will have access to these interim results and make the final decision to terminate the trial | 12 |

| | | | |
|---|---|---|---|
| Harms | 22 | Plans for collecting, assessing, reporting, and managing solicited and spontaneously reported adverse events and other unintended effects of trial interventions or trial conduct | Supplement Section 9 |
| Auditing | 23 | Frequency and procedures for auditing trial conduct, if any, and whether the process will be independent from investigators and the sponsor | NA |
| **Ethics and dissemination** | | | |
| Research ethics approval | 24 | Plans for seeking research ethics committee/institutional review board (REC/IRB) approval | Supplement Section 3 |
| Protocol amendments | 25 | Plans for communicating important protocol modifications (eg, changes to eligibility criteria, outcomes, analyses) to relevant parties (eg, investigators, REC/IRBs, trial participants, trial registries, journals, regulators) | Supplement Section 15 |
| Consent or assent | 26a | Who will obtain informed consent or assent from potential trial participants or authorised surrogates, and how (see Item 32) | Supplement Section 3 |
| | 26b | Additional consent provisions for collection and use of participant data and biological specimens in ancillary studies, if applicable | NA |
| Confidentiality | 27 | How personal information about potential and enrolled participants will be collected, shared, and maintained in order to protect confidentiality before, during, and after the trial | Supplement Section 16 |
| Declaration of interests | 28 | Financial and other competing interests for principal investigators for the overall trial and each study site | Title Page |
| Access to data | 29 | Statement of who will have access to the final trial dataset, and disclosure of contractual agreements that limit such access for investigators | Supplement Section 16 |
| Ancillary and post-trial care | 30 | Provisions, if any, for ancillary and post-trial care, and for compensation to those who suffer harm from trial participation | NA |
| Dissemination policy | 31a | Plans for investigators and sponsor to communicate trial results to participants, healthcare professionals, the public, and other relevant groups (eg, via publication, reporting in results databases, or other data sharing arrangements), including any publication restrictions | Supplement Section 4 |

| | | | |
|---|---|---|---|
| | 31b | Authorship eligibility guidelines and any intended use of professional writers | NA |
| | 31c | Plans, if any, for granting public access to the full protocol, participant-level dataset, and statistical code | Supplement Sections 15-16 |
| **Appendices** | | | |
| Informed consent materials | 32 | Model consent form and other related documentation given to participants and authorized surrogates | NA |
| Biological specimens | 33 | Plans for collection, laboratory evaluation, and storage of biological specimens for genetic or molecular analysis in the current trial and for future use in ancillary studies, if applicable | NA |

*It is strongly recommended that this checklist be read in conjunction with the SPIRIT 2013 Explanation & Elaboration for important clarification on the items. Amendments to the protocol should be tracked and dated. The SPIRIT checklist is copyrighted by the SPIRIT Group under the Creative Commons "Attribution-NonCommercial-NoDerivs 3.0 Unported" license.

#### 3. Ethics and Regulatory Approval

Critically ill patients undergoing tracheal intubation are at significant risk for morbidity and mortality from their underlying illness. Most patients undergoing tracheal intubation in routine clinical care are intubated using either a hyperangulated blade or a standard-geometry blade. Any benefits or risks of these two approaches are experienced by patients undergoing tracheal intubation in clinical care, outside the context of research. As a requirement for enrollment in the ANGLE trial, the patient's treating clinician must determine that either a hyperangulated blade or a standard-geometry blade would be a safe and reasonable approach for that patient. Otherwise, the patient is excluded. Therefore, making the decision between the two approaches randomly in the context of a trial, rather than by a clinician who thinks either approach is safe and reasonable for the patient, is expected to pose no more than minimal additional risk.

Obtaining informed consent for participation in the trial would be impracticable. Most patients undergoing emergency tracheal intubation lack decisional capacity because of their critical illness and surrogate decision-makers are frequently unavailable. Further, emergency tracheal intubation is a time-sensitive procedure with only minutes between the decision to intubate and the completion of the procedure. Meaningful informed consent could not be executed in this brief window.

Because the trial involves minimal risk, the trial would not adversely affect the welfare or privacy rights of the participant, and obtaining informed consent would be impracticable, a waiver of informed consent was requested from and approved by the single institutional review board (IRB) at Vanderbilt University Medical Center (Identifier: 250433). Conduct of the trial with waiver of informed consent is consistent with previous randomized trials comparing alternative approaches to tracheal intubation in widespread clinical use.[1–4] IRBs at participating sites reviewed the protocol, addressed any local contextual factors, and ceded responsibility for ethics approval and trial oversight to the single IRB that approved the trial under a waiver of informed consent.

Information regarding the trial is provided using an IRB-approved patient and family information sheet that clinicians or research personnel give to patients and families, place with patient possessions, or leave at the patient's bedside. The patient and family information sheet contains information on the purpose of the trial, trial procedures, risks and discomforts, benefits, use of protected health information, confidentiality, and investigator contact information.

#### 4. Patient and Public Involvement

The design and conduct of the ANGLE trial—including its integration into clinical care, enrollment of patients unable to provide prospective informed consent, post-enrollment notification, and patient-facing materials—were informed by more than a decade of engagement with patients, families, and community members across more than 10 randomized trials of emergency airway management.[1,2,4–12] People with lived experience of critical illness and emergency tracheal intubation have served as patient investigators, patient partners, and trial steering committee members. Engagement has also included Community Engagement Studios; town halls and meetings with community organizations; surveys conducted online and in ED and ICU waiting rooms; trial websites; and campaigns to disclose trial information and disseminate results. In the most recently published airway management trial conducted by this group, community consultation engaged more than 1,000 people, including 789 surveyed in ED and ICU waiting rooms, and more than 200 community organizations were invited to participate in town halls or Community Engagement Studios. Social media public-disclosure activities reached more than 1.2 million people, and the trial website (the same website at which information about the ANGLE trial is currently publicly posted) received more than 16,000 unique visitors.

These activities provided substantive input into the design and conduct of emergency airway management trials. Participants advised investigators on the acceptability of enrollment when prospective informed consent is infeasible; what patients and families would want to know about the research; when, how, and in what format they would want to be notified; and how patients could express concerns or preferences regarding participation. Patient partners and Community Engagement Studio participants directly reviewed the content, organization, readability, and lay language of patient information sheets. They also advised investigators on the patient-centeredness of trial outcomes, engagement of communities traditionally underrepresented in research, and communication of results to participants and the public. This cumulative input shaped the enrollment and notification processes and patient-facing materials used in ANGLE.

A survivor of critical illness requiring emergency tracheal intubation serves as a full voting member of the ANGLE Data and Safety Monitoring Board. After completion of the trial, investigators will disseminate the results online and through social media using language and formats accessible to patients, families, and the public.

**5. Group Assignment Sheet (Hyperangulated blade group)**

ANGLE Study ID # ______

### Hyperangulated

Use a **hyperangulated video laryngoscope**
on the first attempt.

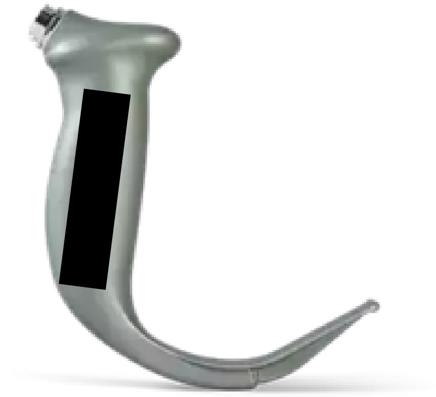

*Examples of hyperangulated video laryngoscopes include the
Storz D-Blade, Glidescope LoPro/Hyperangle S/GVL, and the McGrath X*

If the first attempt fails, use any device or
approach on subsequent attempts.

##### 6. Group Assignment Sheet (Standard-geometry blade group)

ANGLE Study ID # ____

### Standard Geometry

Use a **standard geometry video laryngoscope** on the first attempt.

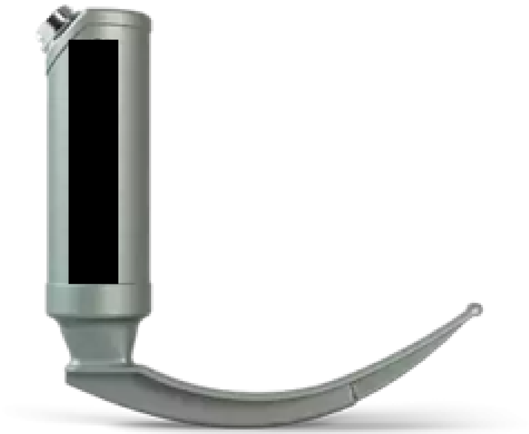

*Examples of standard geometry video laryngoscopes include the Storz C-MAC, Glidescope MAC T/MAC S, and McGrath MAC*

If the first attempt fails, use any device or approach on subsequent attempts.

#### 7. Angle Between the Proximal and Distal Blade Segments of Hyperangulated vs Standard-Geometry Video Laryngoscope Blades

| Manufacturer | Model of Blade | Size | Angle between the proximal and distal segments of the blade (degrees) |
|---|---|---|---|
| **Video Laryngoscope with a Standard-Geometry Blade*** | | | |
| Ambu | SureSight Mac | 3 | 50.5 |
| Ambu | SureSight Mac | 4 | 53.3 |
| Curaplex | OneScope Pro-Direct Mac | 4 | 40.7 |
| Intersurgical | i-view | — | 41.9 |
| Karl Storz | C-MAC Mac† | 3 | 40.4 |
| Karl Storz | C-MAC Mac† | 4 | 47.5 |
| Karl Storz | C-MAC S Mac (single-use) | 4 | 49.4 |
| Medtronic | McGrath MAC† | 3 | 44.7 |
| Medtronic | McGrath MAC† | 4 | 56.1 |
| Verathon | GlideScope Spectrum Mac† | S3 | 50.5 |
| Verathon | GlideScope Spectrum Mac† | S4 | 54.5 |
| **OVERALL** | **--** | **--** | **Median, 49.4 (IQR, 41.9 to 53.3)** |
| **Video Laryngoscope with a Hyperangulated Blade*** | | | |
| Karl Storz | C-MAC D-Blade† | Adult | 79.2 |
| Karl Storz | C-MAC S D-Blade (single-use) | Adult | 75.3 |
| Medtronic | McGrath† | X3 | 80.4 |
| Saturn Biomedical Systems | GlideScope GS-1001-100 | — | 63.3 |
| Verathon | GlideScope GVL Stat† | 3 | 95.6 |
| Verathon | GlideScope GVL Stat† | 4 | 90.2 |
| Verathon | GlideScope Spectrum Hyperangle† | S3 | 62.3 |
| Verathon | GlideScope Spectrum Hyperangle† | S4 | 83.1 |
| **OVERALL** | **--** | **--** | **Median, 79.8 (IQR, 69.3–86.7)** |

*The median angle between the proximal and distal blade segments is 79.8° (interquartile range, 69.3–86.7°) for non-channeled hyperangulated blades and 49.4° (interquartile range, 41.9–53.3°) for standard-geometry video laryngoscope blades. For comparison, the corresponding angles of the Teleflex (Rüsch) Polaris Macintosh direct laryngoscope blades are 44.5° for size 3 and 53.8° for size 4.[13]
†Indicates a blade model available at one or more participating ANGLE trial sites at the time the statistical analysis plan was finalized.

#### 8. Definitions of Free-Day Outcomes

*Ventilator-free days through day 28 (VFDs):* VFDs are defined as the number of calendar days, between enrollment and 28 days after enrollment, on which the patient is alive and free of invasive mechanical ventilation. If a patient is liberated from invasive mechanical ventilation, returns to invasive mechanical ventilation and subsequently is liberated from invasive mechanical ventilation again prior to day 28, the number of VFDs will be counted from the end of the last period of invasive mechanical ventilation to day 28. If the patient is receiving invasive mechanical ventilation at day 28 or dies prior to day 28, VFDs are 0. If a patient is discharged while receiving invasive mechanical ventilation, VFDs are 0. Outcome ascertainment ends at 28 days or hospital discharge, whichever occurs first.

*ICU-free days through day 28 (ICU-FDs):* ICU-FDs are defined as the number of calendar days, between enrollment and 28 days after enrollment, on which the patient is alive and not admitted to an intensive care unit after the patient's final transfer out of the intensive care unit. Patients who are never transferred out of the intensive care unit receive a value of 0. Patients who die before day 28 receive a value of 0. For patients who are transferred out of the ICU, return to an ICU, and are subsequently transferred out of the ICU again prior to day 28, ICU-free days are counted from the date of final transfer out of the ICU. Outcome ascertainment ends at 28 days or hospital discharge, whichever occurs first.

#### 9. Safety Monitoring and Adverse Events

Ensuring patient safety is an essential component of the ANGLE trial. Use of a hyperangulated video laryngoscope blade and use of a standard-geometry video laryngoscope blade are both standard-of-care interventions that have been used in clinical practice for decades with an established safety profile. However, any trial conducted during a high-risk, time-sensitive procedure like tracheal intubation of critically ill patients raises unique safety considerations. The ANGLE trial addresses these considerations through:

1. Exclusion criteria designed to prevent enrollment of patients likely to experience adverse events from intubation using a hyperangulated video laryngoscope blade or intubation using a standard-geometry video laryngoscope blade;
2. Systematic collection of outcomes relevant to the safety of intubation using a hyperangulated video laryngoscope blade or intubation using a standard-geometry video laryngoscope blade;
3. Structured monitoring, assessment, recording, and reporting of adverse events.

##### *9.1 Adverse Event Definitions*

**Adverse Event –** An adverse event is defined as any untoward or unfavorable medical occurrence in a human subject temporally associated with the subject's participation in the research, whether or not considered related to the subject's participation in the research. Any adverse event occurring during the research is classified according to the following characteristics:

- **Seriousness** – An adverse event will be considered "serious" if it:

- Results in death;
- Is life-threatening (defined as placing the patient at immediate risk of death);
- Results in inpatient hospitalization or prolongation of existing hospitalization;
- Results in a persistent or significant disability or incapacity;
- Results in a congenital anomaly or birth defect; or
- Based upon appropriate medical judgment, may jeopardize the patient's health and may require medical or surgical intervention to prevent one of the other outcomes listed in this definition.

● **Unexpectedness** – An adverse event is considered "unexpected" if the nature, severity, or frequency is neither consistent with:
  - The known or foreseeable risk of adverse events associated with the procedures involved in the research that are described in the protocol-related documents, such as the IRB-approved research protocol; nor
  - The expected natural progression of any underlying disease, disorder, or condition of the subject experiencing the adverse event and the subject's predisposing risk factor profile for the adverse event.

● **Relatedness** – The strength of the relationship of an adverse event to a study intervention or study procedure is defined as follows:
  - <u>Definitely Related</u>: The adverse event follows (1) a reasonable, temporal sequence from a study procedure AND (2) cannot be explained by the known characteristics of the patient's clinical state or other therapies AND (3) evaluation of the patient's clinical state indicates to the investigator that the experience is definitely related to study procedures.

- Probably or Possibly Related: The adverse event meets some but not all of the above criteria for “Definitely Related”.
- Probably Not Related: The adverse event occurred while the patient was on the study but can reasonably be explained by the known characteristics of the patient’s clinical state or other therapies.
- Definitely Not Related: The adverse event is definitely produced by the patient’s clinical state or by other modes of therapy administered to the patient.
- Uncertain Relationship: The adverse event does not fit in any of the above categories.

##### *9.2 Monitoring for Adverse Events*

The time interval during which patients are monitored for the occurrence of adverse events begins at randomization and ends at the first of hospital discharge or 28 days. Adverse events occurring before randomization or after hospital discharge or 28 days are not collected. The lead investigator at each enrolling site has primary responsibility for overseeing the monitoring, assessment, and reporting of adverse events. Site study personnel evaluate for the occurrence of adverse events by manual review of the electronic health record at two time points. The first occurs as close as feasible to 24 hours after randomization during initial data collection. The second occurs at the first of hospital discharge or 28 days after enrollment during final data collection. Study personnel at each site communicate regularly with the treating clinicians who perform tracheal intubation in the study environments between enrollment and 28 days after enrollment to solicit information about any potential adverse events. If study personnel at a site identify a potential adverse event, the lead investigator at the site is immediately notified.  The lead investigator at the site assesses the seriousness, unexpectedness, and relatedness of the potential adverse event.  With assistance as needed from the coordinating center and the trial primary investigator, the lead investigator at the site determines whether the event qualifies for recording and reporting.

##### *9.3 Recording and Reporting Adverse Events*

The following types of adverse events are recorded and reported:

- Adverse events that are <u>Serious</u> and <u>Definitely Related, Probably or Possibly Related</u>.
- Adverse events that are <u>Unexpected</u> and <u>Definitely Related, Probably or Possibly Related</u>.

Adverse events that do not meet the above criteria are not recorded or reported. Adverse events that the lead investigator at a site assesses to meet the above criteria for recording and reporting will be entered into the adverse event electronic case report form in the trial database. The lead investigator at the site will record an assessment of each characteristic for the adverse event, including seriousness, unexpectedness, and relatedness. For any adverse event that is **serious AND unexpected**, and definitely related, probably or possibly related, the lead investigator at the site reports the adverse event to the coordinating center and the trial primary investigators **within 24 hours** of becoming aware of the adverse event. For any other adverse event requiring recording and reporting, the lead investigator at the site reports the adverse event to the coordinating center and the trial primary investigators **within 72 hours** of becoming aware of the adverse event. The coordinating center and the trial principal investigator coordinates with the lead investigator at the site to obtain information about the adverse event regarding each characteristic for the adverse event, including seriousness, expectedness, and relatedness. The lead investigator at the site is responsible for making final determinations regarding seriousness and unexpectedness. The coordinating center and trial principal investigator is responsible for making final determinations regarding relatedness.

For adverse events that meet the above criteria for recording and reporting, the coordinating center notifies the DSMB and the IRB in accordance with the following reporting plan:

| **Characteristics of the Adverse Event** | **Reporting Period** |
|---|---|
| Fatal or life-threatening (and therefore serious), unexpected, and definitely related, probably or possibly related. | Report to the DSMB and IRB within 7 days after notification of the event. |
| Serious but non-fatal and non-life-threatening, unexpected, and definitely related, probably or possibly related. | Report to DSMB and IRB within 15 days of notification of the event. |
| All other adverse events meeting criteria for recording and reporting. | Report to DSMB in regularly scheduled DSMB safety reports. |

The coordinating center distributes the written summary of the DSMB's periodic review of reported adverse events to the IRB and study funder.

##### *9.4 Clinical Outcomes that may be Exempt from Adverse Event Recording and Reporting*

In this study of critically ill patients at high risk for death and other adverse outcomes due to their underlying critical illness, clinical outcomes, including death and organ dysfunction, are systematically collected and analyzed for all patients. The primary, safety, and exploratory outcomes are recorded and reported as clinical outcomes and not as adverse events unless treating clinicians or site investigators believe the event is Definitely Related or Probably or Possibly Related to the study intervention or study procedures. This approach – considering death and organ dysfunction as clinical outcomes rather than adverse events and systematically collecting these clinical outcomes for analysis – is common in ICU trials. This approach ensures comprehensive data on death and organ dysfunction for all patients, rather than relying on sporadic adverse event reporting to identify these important events. The following events are examples of study-specific clinical outcomes that would not be recorded and reported as adverse events unless treating clinicians or site investigators believe the event was Definitely Related or Probably or Possibly Related to the study intervention or study procedures:

- Intubation failure
- Cricothyrotomy
- Death (all deaths occurring prior to hospital discharge or 28 days will be recorded);
- Organ dysfunction
  - Pulmonary – hypoxemia, aspiration, acute hypoxemic respiratory failure
  - Cardiac – hypotension, shock, vasopressor receipt, cardiac arrest;
- Duration of mechanical ventilation;
- Duration of ICU admission;
- Duration of hospitalization

#### 10. Composition and Responsibilities of the Data and Safety Monitoring Board

The Data and Safety Monitoring Board (DSMB) consists of members with expertise in bioethics, emergency medicine, pulmonary and critical care medicine, anesthesia, biostatistics, and clinical trials. One patient partner who is a survivor of critical illness requiring emergency tracheal intubation serves as a member of the DSMB. All members of the DSMB are voting members. The DSMB developed a charter, approved the trial protocol, approved the patient notification forms, and approved the trial monitoring plan prior to trial initiation of enrollment. The DSMB has the ability at any point to recommend that the trial end, be modified, or continue unchanged.

The principal role of the DSMB is to assure the safety of patients in the trial. The DSMB monitors data from the trial at semi-annual meetings, reviews and assesses the performance of its operations, and makes recommendations to the investigators with respect to:

- Review of adverse events
- Interim results of the study for evidence of efficacy or adverse events
- Possible early termination of the trial because of new external information, early attainment of study objectives, safety concerns, or inadequate performance
- Possible modifications in the clinical trial protocol
- Performance of individual centers

#### 11. Additional Analyses of the Primary Outcome

We will assess the robustness of the findings of the primary analysis in a series of sensitivity analyses using different approaches to defining and analyzing the primary outcome. First, we will repeat the primary analysis among patients whose primary outcome was assessed by an independent observer, excluding patients for whom operator self-report was the sole source of primary outcome data. Second, we will repeat the primary analysis adjudicating all patients whose first laryngoscopy attempt was performed with the non-assigned blade (i.e., crossover) as not having successful intubation on the first attempt. Third, because an operator's prior experience with each blade geometry may affect the relative effectiveness of hyperangulated and standard-geometry blades, we will repeat the primary analysis among patients whose operator reported a similar amount of prior experience using both hyperangulated and standard-geometry blades (i.e., using a hyperangulated blade for 25% to 75% of their prior intubations with a video laryngoscope). Fourth, we will perform an adjusted analysis comparing the primary outcome between groups using a generalized linear mixed effects model with a random effect for trial site and fixed effects for trial group assignment and the following pre-specified baseline variables: age, sex, body mass index, the operator's pre-enrollment assessment of the anticipated difficulty of intubation (easy, moderate-or-difficult, or not recorded), the presence of cervical spine immobilization, the operator's pre-enrollment assessment of the presence of any difficult airway characteristics, operator experience quantified as the operator's total number of previous intubations, and the location in which the intubation procedure was being performed (emergency department vs intensive care unit). Continuous variables will be modeled using restricted cubic splines with 3 to 5 knots to allow for potential nonlinear associations with the outcome.

#### 12. Analyses of heterogeneity of treatment effect

Heterogeneity of treatment effect is nonrandom variation in the magnitude or direction of the effect of a treatment on an outcome across levels of a baseline covariate. We will examine whether prespecified baseline variables modify the effect of trial group assignment (hyperangulated blade vs standard-geometry blade) on the primary outcome using a formal test of statistical interaction in a logistic regression model with the primary outcome as the dependent variable and trial group, the proposed effect modifier, and the interaction between trial group and the proposed effect modifier included as independent variables. For categorical variables, we will present the absolute difference and 95% CIs within each prespecified subgroup. Continuous variables may be dichotomized for presentation. In accordance with the Instrument for assessing the Credibility of Effect Modification Analyses (ICEMAN) recommendations,[14] we have prespecified the following limited number of baseline variables as potential effect modifiers and the hypothesized direction of effect modification for each:

1. Body mass index (kg/m$^2$): We hypothesize that body mass index will modify the effect of trial group assignment on the primary outcome, with a greater increase in the incidence of successful intubation on the first attempt with the use of a hyperangulated blade compared with a standard-geometry blade among patients with higher body mass index as compared with patients with lower body mass index. The hypothesis of effect modification is supported by data from previous randomized trials in the operating room suggesting a greater benefit to use of a hyperangulated blade for patients with higher body mass index.[15–17]
2. Patient location (ED / ICU): We hypothesize that the location of the patient at the time of enrollment will not modify the effect of trial group assignment on the primary outcome.

3. Trauma: We hypothesize that the occurrence of a traumatic injury prior to enrollment will modify the effect of trial group assignment on the primary outcome, with a greater incidence of successful intubation on the first attempt with the use of a hyperangulated blade compared with a standard-geometry blade among patients who have experienced traumatic injury, as compared with those who have not. This hypothesis of effect modification is supported by (i) observational data suggesting a higher incidence of successful intubation on the first attempt with use of a hyperangulated blade and (ii) indirect inference from the results of a randomized trial that demonstrated an increased incidence of successful intubation on the first attempt with use of a video laryngoscope with a hyperangulated blade compared to a direct laryngoscope.[18–20]
4. Cervical spine immobilization: We hypothesize that cervical spine immobilization with a cervical collar or manual in-line stabilization will modify the effect of trial group assignment on the primary outcome, with a greater incidence of successful intubation on the first attempt with the use of a hyperangulated blade compared with a standard-geometry blade among patients who have cervical immobilization, as compared with those who have not. This hypothesis of effect modification is supported by evidence of an increased incidence of successful intubation on the first attempt with use of a video laryngoscope with a hyperangulated blade compared to a direct laryngoscope among patients with trauma, for whom cervical spine immobilization has been proposed as the key effect modifier.[18–20]
5. Operator's pre-enrollment assessment of the anticipated difficulty of intubation (Easy, Moderate-or-difficult, Not Recorded): We hypothesize that the operator's pre-enrollment assessment of the anticipated difficulty of intubation will modify the effect of trial group assignment on the primary outcome, with a greater increase in the

incidence of successful intubation on the first attempt with the use of a hyperangulated blade compared with a standard-geometry blade among patients assessed as moderate or difficult, as compared with easy. This hypothesis of effect modification is supported by a randomized trial reporting a higher incidence of successful intubation on the first attempt with use of a hyperangulated blade compared to a standard-geometry blade in patients with an anticipated difficult airway undergoing intubation in the operating room.[21]

6. Operator's pre-enrollment assessment of the presence of difficult airway characteristics (Present, Absent, Not Recorded): We hypothesize that the operator's pre-enrollment assessment of the presence of any difficult airway characteristics (i.e., small or recessed chin, short neck, large neck circumference, limited range of motion in neck, cervical collar or in-line stabilization) will modify the effect of trial group assignment on the primary outcome, with a greater increase in the incidence of successful intubation on the first attempt with the use of a hyperangulated blade compared with a standard-geometry blade among patients assessed as having one or more difficult airway characteristics. This hypothesis of effect modification is supported by a randomized trial reporting a higher incidence of successful intubation on the first attempt with use of a hyperangulated blade compared to a standard-geometry blade in patients with an anticipated difficult airway undergoing intubation in the operating room.[21]
7. Operator experience at the time of enrollment:
    a. Total number of operator's previous intubations: We hypothesize that the total number of previous intubations performed by the operator will modify the effect of trial group assignment on the primary outcome, with a greater increase in the incidence of successful intubation on the first attempt with the

use of a hyperangulated blade compared with a standard-geometry blade among patients intubated by an operator with fewer previous intubations. The hypothesis of effect modification is supported by a randomized trial among critically ill patients showing greater benefit from use of a video laryngoscope among inexperienced operators and a trial in the operating room in which there was an increased incidence of successful intubation on the first attempt with the use of a hyperangulated blade among less experienced operators.[10,15]

b. Proportion of previous intubations with a video laryngoscope performed with a hyperangulated blade. We hypothesize that the proportion of an operator's previous intubations performed using a hyperangulated blade (rather than a standard-geometry blade) will modify the effect of trial group assignment on the primary outcome, with a greater increase in the incidence of successful intubation on the first attempt with use of a hyperangulated blade compared with a standard-geometry blade among operators with a higher proportion of the operator's previous intubations performed using a hyperangulated blade. The hypothesis of effect modification is supported by an individualized treatment effect analysis derived from a prior randomized trial evaluating procedural tools and techniques that demonstrated an increased incidence of successful intubation on the first attempt with a bougie among operators with more prior experience using a bougie.[3,22]

We will also perform an effect modification analysis for the primary outcome that includes a three-way interaction between trial group, total number of previous intubations performed by the operator, and proportion of previous intubations performed by the operator using a hyperangulated blade.

#### 13. Rationale for the approach to collecting, reporting, and analyzing difficult airway characteristics and anticipated difficulty of intubation

In the ANGLE trial, after the decision has been made to intubate, but before the envelope has been opened, the clinicians who will be performing the intubation procedure record on a structured case record two variables related to the anticipated difficulty of performing intubation for the patient. Recording these variables prior to randomization is intended to prevent knowledge of trial group assignment from potentially influencing the clinicians' assessments.

First, clinicians use their knowledge of the patient's anatomy, physiology, and clinical course to make a global assessment of the expected difficulty of tracheal intubation (easy, moderate, difficult). This same subjective global assessment was collected prior to enrollment in a previous trial for 1417 patients undergoing tracheal intubation in the same ED and ICU settings as the ANGLE trial. In that study, the subjective global assessment appeared to discriminate between who went on to experience successful intubation on the first attempt vs those who did not: the incidence of successful intubation on the first attempt for patients in the video laryngoscope group of the trial was 88.8% for patients anticipated by clinicians to be "easy", 83.9% for patients anticipated to be "moderate", and 76.1% for patients anticipated to be difficult.[10] These data suggest that clinicians' subjective assessment appears to perform well as a baseline risk assessment for the primary outcome of the ANGLE trial in the settings in which the ANGLE trial is being conducted.

Second, clinicians record the presence or absence of each of the following pre-specified difficult airway characteristics: small or recessed chin; short neck; large neck circumference; limited range of motion in neck; and cervical collar or in-line stabilization. These difficult airway characteristics were selected by the investigators during the design of the trial with input from emergency medicine, anesthesiology, and critical care clinicians as (i) objective characteristics that (ii) are common in the patient population for the trial, (iii) can be assessed prior to initiation of the laryngoscopy, and (iv) would be hypothesized to modify the effect of use of hyperangulated vs standard-geometry blade on the outcome of successful intubation on the first attempt.

The ANGLE trial collected (i) this global subjective assessment of the anticipated difficulty of intubation and (ii) this prespecified set of difficult-airway characteristics rather than a formal difficult airway prediction score, such as the MACOCHA score.[23,24] The 7-component MACOCHA score uses characteristics of the patient (Mallampati score, obstructive sleep apnea, reduced mobility of cervical spine, limited mouth opening), the patient’s illness (severe hypoxia, coma), and the operator (non-anesthesiologist) to predict ICU patients’ risk of difficult intubation, defined as more than three laryngoscopy attempts or intubation lasting more than 10 minutes. The MACOCHA score was derived and validated in French ICUs.[23] Prior to the design of the ANGLE trial, we evaluated the performance of the MACOCHA score for predicting successful intubation on the first attempt in ICUs in the United States that would go on to participate in the ANGLE trial. We found that, among 130 critically ill adults undergoing tracheal intubation in five intensive care units (ICUs) during the Checklists and Upright Positioning in Endotracheal Intubation (Check-UP) trial,[6] the MACOCHA score’s area under the curve (AUC) for the receiver operating characteristic curve (ROC) for the outcome of successful intubation on the first attempt was 0.58 (95% CI, 0.47 to 0.70). Thus, using the MACOCHA score in these settings to predict successful intubation on the first attempt was not significantly better than chance alone.

Moreover, a main goal of collecting information on the anticipated difficulty of intubation in the ANGLE trial is to evaluate whether that baseline patient characteristic modifies the effect of a hyperangulated blade vs standard-geometry blade on successful intubation on the first attempt. The MACOCHA score was not designed for this use. The MACOCHA score combines features that might make use of a hyperangulated blade more beneficial (e.g., cervical spine immobilization) and other features that would not be expected to make use of a hyperangulated blade more beneficial (e.g., presence of hypoxemia at the time of induction).

#### 14. Rationale for the approach to collecting, reporting, and analyzing operator experience

To account for the operator's overall and blade-specific experience, immediately after the intubation procedure the operator completes a paper data collection form to record characteristics of the procedure. This includes the operator's name, specialty (e.g., emergency medicine, critical care, anesthesia), and level of training (e.g., resident, fellow, attending, certified registered nurse anesthetist, nurse practitioner, physician assistant). The operator records the total number of intubations performed using a direct laryngoscope, the total number of intubations performed using a video laryngoscope with a hyperangulated blade, and the total number of performed intubations using a video laryngoscope with a standard-geometry blade. This level of detail will allow us to report the operator's range of overall experience and blade-specific experience and evaluate for heterogeneity of treatment effect by overall experience and blade-specific experience.

#### 15. Plan for Communication of Protocol Changes

Any changes to the trial protocol, including changes to eligibility, outcomes, and analyses, will be implemented via a new version of the full trial protocol, tracked with the date of the update, and the version number of the trial protocol. A list summarizing the changes made with each protocol revision will be included at the end of each protocol. The updated protocol will be sent to the sIRB for approval and tracking prior to implementation of the protocol change. At the time of publication, the original trial protocol and the final trial protocol, including the summary of changes made with each protocol version, will be included in the supplementary material for publication.

#### 16. Patient Privacy, Data Storage, and Sharing

Federal regulations 45 CFR 46.111 (a) (7) requires that, when appropriate, there are adequate provisions to protect the privacy of patients and to maintain the confidentiality of data. At no time during the course of this trial, its analysis, or its publication will patient identities be revealed in any manner. The minimum necessary data containing patient or provider identities are collected in ANGLE. All patients are assigned a unique study ID number for tracking. The PHI required to accurately collect clinical and outcomes data is available only to investigators at the site at which the subject is enrolled, and these data are shared only in completely de-identified form with the coordinating center via the secure online database REDCap. The de-identified dataset housed in REDCap will be accessed by the coordinating center for reporting the results of this trial. All data will be maintained in the secure online database REDCap until the time of trial publication. At the time of publication, all PHI will be expunged and only the de-identified version of the database will be retained. Potential future use of de-identified data generated in the course of this trial by the coordinating center and other participating sites is permitted as allowed by mutual data sharing use agreements.

#### 17. Trial Status

The ANGLE trial is a pragmatic, multicenter, unblinded randomized clinical trial evaluating the effect of use of a hyperangulated video laryngoscope blade versus use of a standard-geometry video laryngoscope blade on the incidence of successful intubation on the first attempt among critically ill adults undergoing tracheal intubation. Enrollment began on December 30, 2025, and is expected to conclude in late 2026 or early 2027.
